# Analytical and Clinical Validation of AIM-NASH: A Digital Pathology Tool for Artificial Intelligence-based Measurement of Nonalcoholic Steatohepatitis Histology

**DOI:** 10.1101/2024.05.29.24308109

**Authors:** Hanna Pulaski, Stephen A. Harrison, Shraddha S. Mehta, Arun J Sanyal, Marlena C. Vitali, Laryssa C. Manigat, Hypatia Hou, Susan P. Madasu Christudoss, Sara M. Hoffman, Adam Stanford-Moore, Robert Egger, Jonathan Glickman, Murray Resnick, Neel Patel, Cristin E. Taylor, Robert P. Myers, Chuhan Chung, Scott D. Patterson, Anne-Sophie Sejling, Anne Minnich, Vipul Baxi, G. Mani Subramaniam, Quentin M. Anstee, Rohit Loomba, Vlad Ratziu, Michael C Montalto, Andrew H Beck, Katy Wack

**Author notes:** Corresponding author: Katy Wack. Co-first authors (Hanna Pulaski, Stephen A. Harrison). Employee of PathAI at the time of the study (Shraddha S. Mehta, Marlena C. Vitali, Laryssa C. Manigat, Susan P. Madasu Christudoss, Sara M. Hoffman, Murray Resnick, Cristin E. Taylor, Michael C Montalto).

## Abstract

Metabolic-dysfunction associated steatohepatitis (MASH) is a major cause of liver-related morbidity and mortality, yet treatment options are limited. Manual scoring of liver biopsies, currently the gold standard for clinical trial enrollment and endpoint assessment, suffers from high reader variability. This study represents the most comprehensive multi-site analytical and clinical validation of an AI-based pathology system, Artificial Intelligence-based Measurement of Nonalcoholic Steatohepatitis (AIM-NASH), to assist pathologists in MASH trial histology scoring. AIM-NASH demonstrated high repeatability and reproducibility compared to manual scoring. AIM-NASH-assisted reads by expert MASH pathologists were superior to unassisted reads in accurately assessing inflammation, ballooning, NAS >= 4 with >=1 in each score category, and MASH resolution, while maintaining non-inferiority in steatosis and fibrosis assessment. These findings suggest AIM-NASH could mitigate reader variability, providing a more reliable assessment of therapeutics in MASH clinical trials.

## Main

Metabolic dysfunction-associated steatotic liver disease (MASLD), formerly termed nonalcoholic fatty liver disease (NAFLD) [1] is emerging as a significant global health challenge, affecting approximately a quarter of the global population [2]. The progression of MASLD to Metabolic dysfunction-associated steatohepatitis (MASH), formerly known as nonalcoholic steatohepatitis (NASH) has emerged as the foremost reason for liver transplants among women [3], with predictions suggesting it may soon account for the leading overall cause of liver transplant [4]. The urgency of the situation is underscored by the limited number of approved therapeutic interventions by the Food and Drug Administration (FDA) and European Medicines Agency (EMA) for MASH, even though it affects a significant number of patients worldwide. The landscape of drug development in this domain is fraught with trials that have shown borderline results or outright failures based on liver histology.

The challenge is exacerbated by the absence of a broadly reliable and validated histologic scoring mechanism to ascertain patient suitability for clinical trials and to evaluate the success of experimental treatments. Histologic-based assessment of liver biopsies is currently the gold standard for MASH diagnosis. This diagnosis is based on the presence of specific histologic patterns observed in the absence of significant alcohol consumption, and the patterns with extent of fibrosis play a pivotal role in disease staging. The FDA has recognized that alterations in these histologic attributes, observable through liver biopsies, are likely indicative of clinical benefits [5]. Consequently, these score-based disease activity and stage changes are deemed viable surrogate endpoints in MASH clinical trials for accelerated approvals [6, 7]. Key instruments like the MASLD activity score (MAS) by the MASH Clinical Research Network (CRN) facilitate disease activity measurement [8], while the CRN fibrosis scale evaluates fibrosis progression or improvement and is an influential predictor of long-term outcomes [9]. Regulatory bodies such as the FDA and the EMA predominantly rely on the CRN MASH measurement systems to determine surrogate endpoints [6, 7, 10].

The recent emergence of noninvasive tests (NITs) has led to an initiative to replace biopsies with NITs and has been discussed widely in the MASH community. However, there are no NITs or combination of NITs that are currently analytically or clinically validated for broad use in trials, which demonstrate high sensitivity, specificity, and reproducible grading and staging of patients with MASH for use as surrogate endpoints in MASH clinical trials. Acquiring this validation data, including clinical outcomes across multiple drug candidates, will take years. Many biopsy-based MASH clinical trials are currently in phase 2 and approaching phase 3 trials, and the recent accelerated approval of Resmetirom was achieved through consensus scoring and several re-read methods utilized to confirm histologic score-based primary endpoints [11]. This burdensome approach can be necessary to overcome questions around reader bias and variability, which can affect accuracy of histologic-based score change and, therefore, determination of whether a drug candidate has met its primary endpoint or to measure and monitor its efficacy. Additionally, the current gold standard approach has been shown to still be subject to significant inter-panel variability, demonstrating that there is still lack of standardization and therefore of reliable, accurate scoring [12]. Full approval will not occur until clinical outcomes show a favorable benefit to risk profile in treated patients, when these results are collected over multiple years. This significant read variability is a major risk for potentially effective treatments to fail in phase 2b trials which have relatively low sample sizes, and phase 3 trials, or to require very costly and burdensome, multiple read strategies to confirm and measure efficacy. Therefore, there is still an urgent unmet need for a tool that can be used by pathologists to enroll and measure histologic change for accelerated approval accurately, precisely, and in a standardized manner. In addition, it will be important to understand the relationship between histologic-based assessment in validating NITs for diagnostic contexts of use.

The interpretation of the current scoring systems present significant challenges to clinical trial outcome analysis, particularly concerning reproducibility [13–17]. Given that the gold standard endpoint for accelerated approval is a difference in histological scores from baseline to treatment timepoints, inherent intra and inter-reader variability can confound the measurement of true drug effect [18]. This variability can significantly undermine the power of a study, posing challenges especially in trials with smaller sample sizes, such as phase 1 and 2 trials. To circumvent this limitation, trials are often required to be over-powered, adding cost and time to trials. Such variability likely arises due to discrepancies in feature interpretation, feature heterogeneity within a biopsy sample and the quantification of these features using scoring systems [19]. Additionally, the current scoring criteria were not developed to quantify change in disease activity.

The rapidly evolving field of artificial intelligence (AI) offers a promising avenue to address these challenges. AI has demonstrated significant advancements in numerous medical disciplines, with a marked rise in CE marked (a standard for European health, safety, performance and environmental requirements) and FDA approved in vitro diagnostics for AI-based medical devices and algorithms from 2015 to 2020 [20] and the FDA approval for an AI product in digital pathology in 2021 for Paige Prostate [21]. However, the field of quantitative pathology in MASH therapeutic development still awaits a tool that is scalable, reproducible, and validated. Recently, we described the development and verification of the AIM-NASH (Artificial Intelligence-based Measurement of Nonalcoholic Steatohepatitis), AI-based clinical trial tool [22]. In this previous body of work, the algorithm was developed and verified (without any pathologist review) for accuracy compared to a panel of manual readers to confirm that the tool was ready to be locked. As proof of concept, the algorithm alone (without pathologist review) was also retrospectively deployed on ATLAS clinical trial dataset to demonstrate the utility of the tool. The work presented here represents extensive, multi-site analytical and clinical validation of the algorithm alone and as an assist to MASH pathologists, as it would be used prospectively in a clinical trial, with each histologic component score being assessed individually and as a part of histologic-based composite inclusion criteria and endpoint determination. This validation study, the largest known of its kind, included approximately 13,000 independent reads for over 1400 biopsies across 4 completed, global MASH clinical trials with various drug mechanisms of action. The study was performed across multiple sites (internal and external) and included samples with extensive variation in disease activity as well as biopsy, staining and scanning quality. Multiple, prospectively collected pathologist reads per case (where readers were either unassisted or assisted by AI) were collected from MASH expert pathologists, including reads from an independent “gold standard” consensus group. These reads were used to externally and robustly test both the algorithm alone and as used as an aid to pathologists (**Figure 1A**), in representative trial settings. This extensive collection of AIM-NASH validation studies and analyses was designed in partnership with FDA, EMA, and multiple experts from academia and drug development over several years of collaborative work. The aim was to demonstrate the tool’s ability to provide a reliable, efficient solution for pathologists to address the urgent unmet need for accurate, standardized, clinical trial enrollment and histologic endpoint assessments, paving the way for more streamlined MASH drug approval pathways.

**Figure 1.**
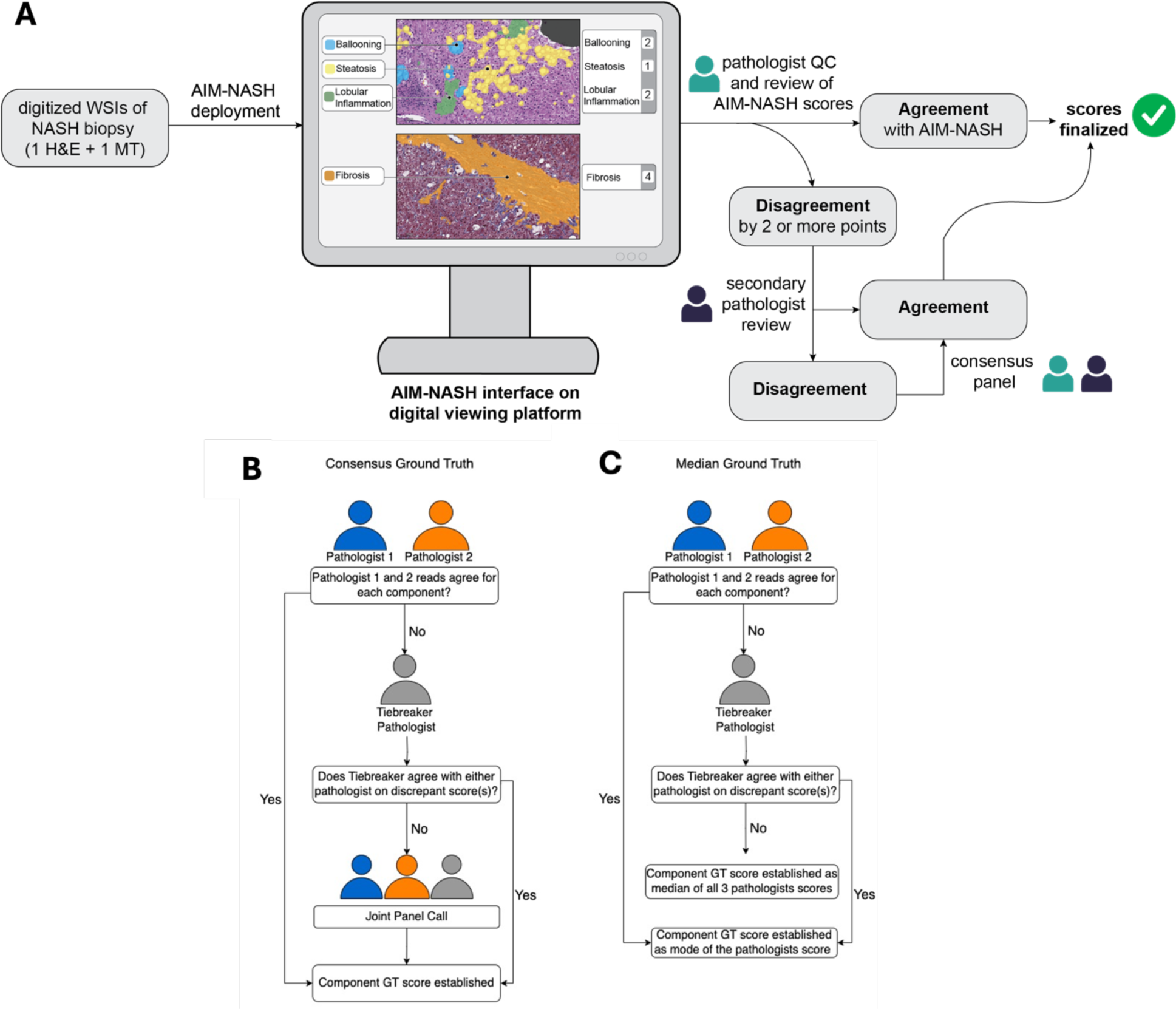
AI-assisted workflow with representative AIM-NASH overlays and ground truth panel workflows. **(A)** In the AI-assisted workflow, the primary pathologist reviews the AIM-NASH output and does a quality control review of the slide (determines if re-stain or rescan of the slide is necessary, confirms all trial specific criteria is met and notes any additional findings). If the primary pathologist disagrees with any MASH component(s) by 2 points or more, the case goes to a review by a secondary pathologist, who independently reviews the discordant AIM-NASH score(s). If the secondary pathologist agrees with the primary pathologist’s modified score, this will be the final score, if they disagree with the primary pathologist or agree with AIM-NASH, the 2 pathologists will convene on a consensus call where they agree on the final score. **(B)** Consensus Ground truth (GT) was determined by two panels of hepatopathologists. Each panel consisted of two main reader pathologists and an auxiliary tie-breaker pathologist. Discrepancies in scoring among the primary readers prompted the intervention of the tiebreaker pathologist, who was blind to initial assessments. When the tiebreaker’s scoring diverged from both primary readers, a panel discussion was convened for consensus, with the tiebreaker’s score being decisive in rare cases of continued disagreement. **(C)** For median GT score, when the tiebreaker’s scoring diverged from both primary readers, the median of the 3 scores was considered final. Overall, five distinct pathologists contributed to establishing the GT.

Once a tool such as this is analytically and clinically validated and is fully qualified by FDA and EMA in the Drug Development Tool (DDT) and Novel Methodologies for Drug Development programs, it is then more broadly available for use by pathologists in place of manual scoring for all histologic assessments in MASH trials.

## Results

### Overlay Validation Analyses

Up to 160 frames per feature (steatosis, lobular inflammation, hepatocellular ballooning, fibrosis, H&E artifact, and trichrome artifact) were evaluated in this study (some frames were enrolled for multiple features). Distribution of frames based on slide level score (GT scores) are listed in Table S1 and distribution of frames based on frame level scores (collected from the enrollment pathologist) are listed in Table S2. For each frame and each feature, the pathologists indicated whether the feature was present (yes/no), shown in Table S3.

The acceptance criteria for true positive (TP; underestimation) success were met for all feature overlays except for hepatocellular ballooning, where it was narrowly missed, and the mean success rates were all above 0.85. H&E artifact TP success rate was 0.97 (95% CI, 0.95, 0.99), trichrome artifact was 0.99 (95% CI, 0.97, 1), lobular inflammation was 0.94 (95% CI, 0.92, 0.96), steatosis 0.96 (95% CI, 0.93, 0.98) and fibrosis 0.97 (95% CI, 0.95, 0.99). For hepatocellular ballooning the overall TP success rate was 0.87, with 95% CI (0.83,0.91). The acceptance criteria for false positive (FP; overestimation) success rate were met for all 6 feature overlays. H&E artifact success rate for FP was 0.97 (95% CI, 0.95, 0.99), trichrome artifact was 0.93 (95% CI, 0.90, 0.96), lobular inflammation was 0.99 (95% CI, 0.98, 0.99), steatosis was 1.00 (95% CI, 0.98, 1), hepatocellular ballooning was 0.92 (95% CI, 0.90, 0.94) and fibrosis was 0.99 (95% CI, 0.99, 1).

The individual pathologist TP and FP success rates are listed in Table 1. Proportion of frames where all 3 evaluating pathologists agreed on presence of the feature when at least 1 pathologist indicated presence of feature in a frame was 89% (132 frames out of 148 frames) for H&E artifact, 55.1% for hepatocellular ballooning (65 of 118 frames), 80.0% (124 of 155 frames) for lobular inflammation, 99.4% (1358 of 159 frames), 72.0% (108 of 150 frames) for trichrome artifact and 96.8* (149 of 154 frames). Given that the agreement for presence of hepatocellular ballooning was the lowest between pathologists (55.1%) out of all features and the TP success rate for ballooning was above 0.90 for 2 out of 3 of the pathologists, the sources of variability between pathologists for hepatocellular ballooning were further examined. For the 65 frames where all 3 evaluating pathologists indicated presence of hepatocellular ballooning, the TP success rate was calculated. Pathologists A and B identified underestimation in 1 and 3 of the 65 frames, respectively, resulting in TP success rates of 0.99 for pathologist A and 0.95 for pathologist B for those frames. However, given that pathologist C identified underestimation in 10 of the 65 frames, showing a TP success rate of 0.85, and pathologist C identified a total of 111 frames that had some ballooned cells compared to 92 and 71 for pathologist A and B (Table S3), this indicates that pathologist C may be identifying more cells as ballooned hepatocytes than the other two pathologists and the algorithm. This is expected given the lack of standardization across expert pathologists in both identifying and quantifying ballooned hepatocytes [23].

**Table 1:** True Positive (TP) and False Positive (FP) Success Rate per Individual Pathologist for Overlay Validation.

| Feature | Pathologist | TP Success Rate<br>(95% CI) | FP Success Rate<br>(95% CI) |
| --- | --- | --- | --- |
| H&E Artifact | A | 0.97 (0.94, 0.99) | 0.96 (0.92, 0.99) |
|  | B | 0.98 (0.95, 1.0) | 0.98 (0.95, 0.99) |
|  | C | 0.97 (0.94, 0.99) | 0.99 (0.97, 1.0) |
| Hepatocellular ballooning | A | 0.96 (0.91, 0.99) | 1.00 (0.98, 1.0) |
|  | B | 0.94 (0.89, 0.99) | 1.00 (0.98, 1.0) |
|  | C | 0.72 (0.64, 0.81) | 0.76 (0.70, 0.83) |
| Lobular inflammation | A | 0.98 (0.95, 1.0) | 1.00 (0.98, 1.0) |
|  | B | 0.98 (0.95, 1.0) | 1.00 (0.98, 1.0) |
|  | C | 0.86 (0.81, 0.92) | 0.98 (0.95, 0.99) |
| Steatosis | A | 0.94 (0.90, 0.98) | 1.00 (0.98, 1.0) |
|  | B | 0.99 (0.97, 1.0) | 1.00 (0.98, 1.0) |
|  | C | 0.94 (0.90, 0.98) | 1.00 (0.98, 1.0) |
| Trichrome Artifact | A | 0.99 (0.97, 1.0) | 0.88 (0.82, 0.92) |
|  | B | 0.98 (0.96, 1.0) | 0.94 (0.91, 0.98) |
|  | C | 0.99 (0.97, 1.0) | 0.98 (0.95, 0.99) |
| Fibrosis | A | 0.97 (0.94, 0.99) | 1.00 (0.98, 1.0) |
|  | B | 0.99 (0.98, 1.0) | 1.00 (0.98, 1.0) |
|  | C | 0.95 (0.91, 0.98) | 0.99 (0.98, 1.0) |

### Algorithm Repeatability and Reproducibility

For inter-day scanner repeatability (AIM-NASH deployment on the same glass slides on different scans from the same scanner on different days), mean agreement rates between the AIM-NASH scoring on the 3 separate Whole Slide Images (WSIs) for steatosis was 0.93 (95% CI of (0.89, 0.96), p<0.0001), lobular inflammation was 0.96 (95% CI of (0.94, 0.99), p<0.0001), hepatocellular ballooning was 0.96 (95% CI of (0.93, 0.98), p<0.0001) and fibrosis was 0.93 (95% CI of (0.89, 0.96), p<0.001), (**Figure 2A**).

**Figure 2.**
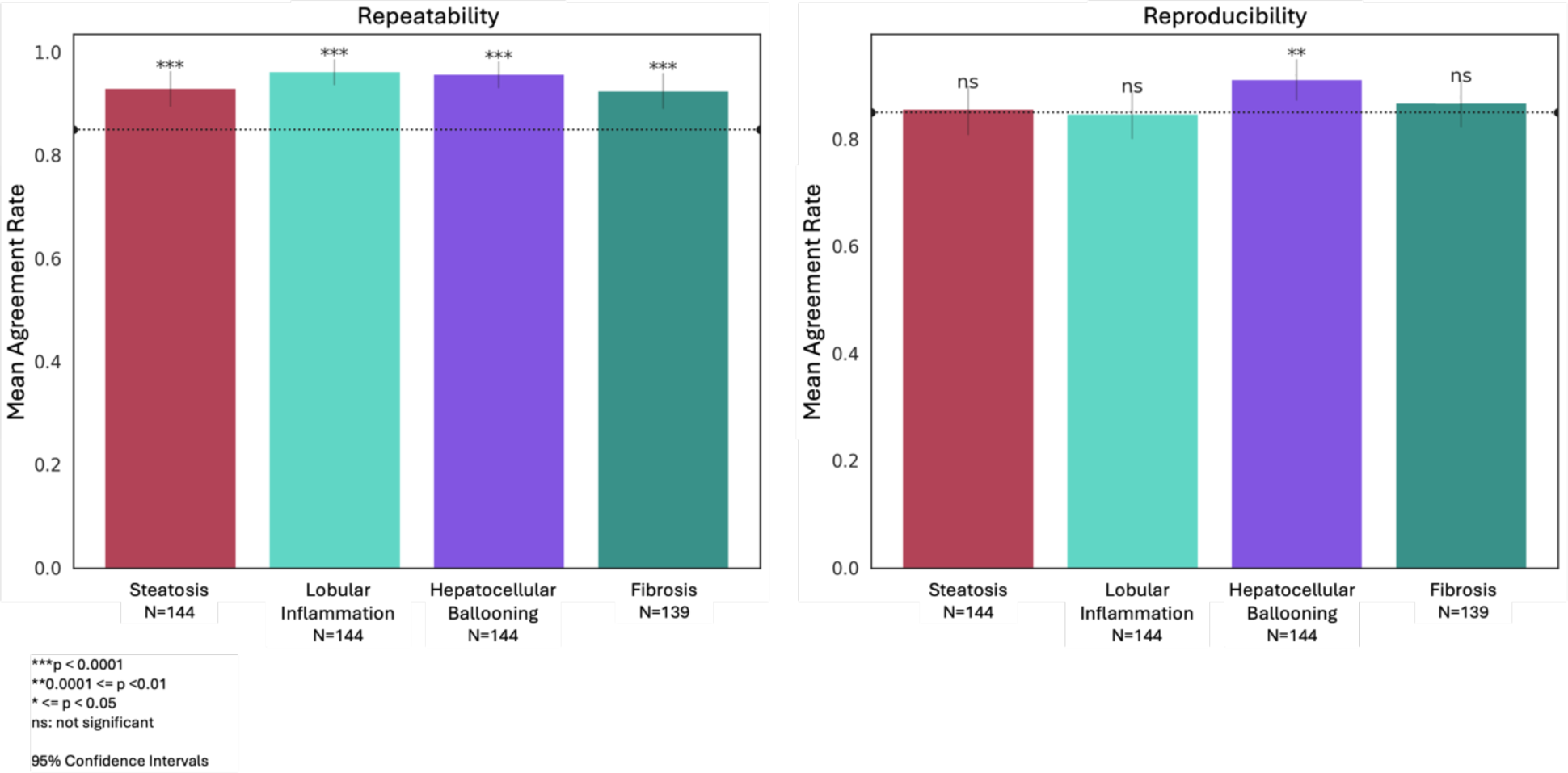
Scanner repeatability and reproducibility of AIM-NASH. **(A)** For scanner repeatability, a subset of 150 cases from the clinical validation were scanned multiple times using the same Leica Aperio AT2 scanner at 40x magnification on 3 non-consecutive days. **(B)** For scanner reproducibility, the same slides were scanned once at 3 different labs by 3 different operators using 3 different Leica Aperio AT2 scanners at 40x magnification.

For inter-site scanner reproducibility (AIM-NASH deployment on the same glass slides on different scans from 3 different sites), mean agreement rate for hepatocellular ballooning was 0.91 (95% CI of 0.87, 0.95, p=0.02), meeting the acceptance criteria. The mean agreement rates for steatosis, lobular inflammation, and fibrosis were approximately 85% but the CIs fell slightly below the 0.85 acceptance criteria (steatosis 0.86 (95% CI of 0.81, 0.9, p=0.39), lobular inflammation 0.85 (95% CI of 0.80, 0.89, p=0.53), and fibrosis 0.87(95% CI of 0.82, 0.91, p=0.21) (**Figure 2B**).

Pairwise inter-reader agreements were calculated between IMR pathologists across all cases (Table S4) to explicitly compare reproducibility across study pathologists to reproducibility achieved by AIM-NASH across sites and scanners. For all histologic components, inter-scan, intra-site repeatability, and inter-scan, inter-site reproducibility was higher than for pathologist mean pairwise agreement (for pairs of pathologists who read at least 10 common cases) (Table 2).

**Table 2.** Manual pathologist vs. AIM-NASH repeatability and reproducibility.

| Feature | Mean AIM-NASH<br>Inter-scan, Intra-site<br>Repeatability<br>(% Agreement) | Mean AIM-NASH<br>Inter-site Reproducibility<br>(% Agreement) | Mean pairwise<br>Agreement for<br>Pathologists<br>(% Agreement) |
| --- | --- | --- | --- |
| Steatosis | 93.1 | 85.6 | 70.3 |
| Lobular inflammation | 95.8 | 84.7 | 45.3 |
| Hepatocellular ballooning | 96.3 | 91.2 | 55.6 |
| Fibrosis | 92.6 | 86.8 | 61.5 |

### Accuracy of Algorithm Alone and as an Assist Tool for Pathologists

Evaluation for non-inferior accuracy of AIM-NASH (algorithm only and AI-assisted) to IMRs was assessed by comparing the mean Weighted Kappa (WK) of IMRs with GT to the WK of AIM-NASH with GT (**Figure 3**).

**Figure 3.**
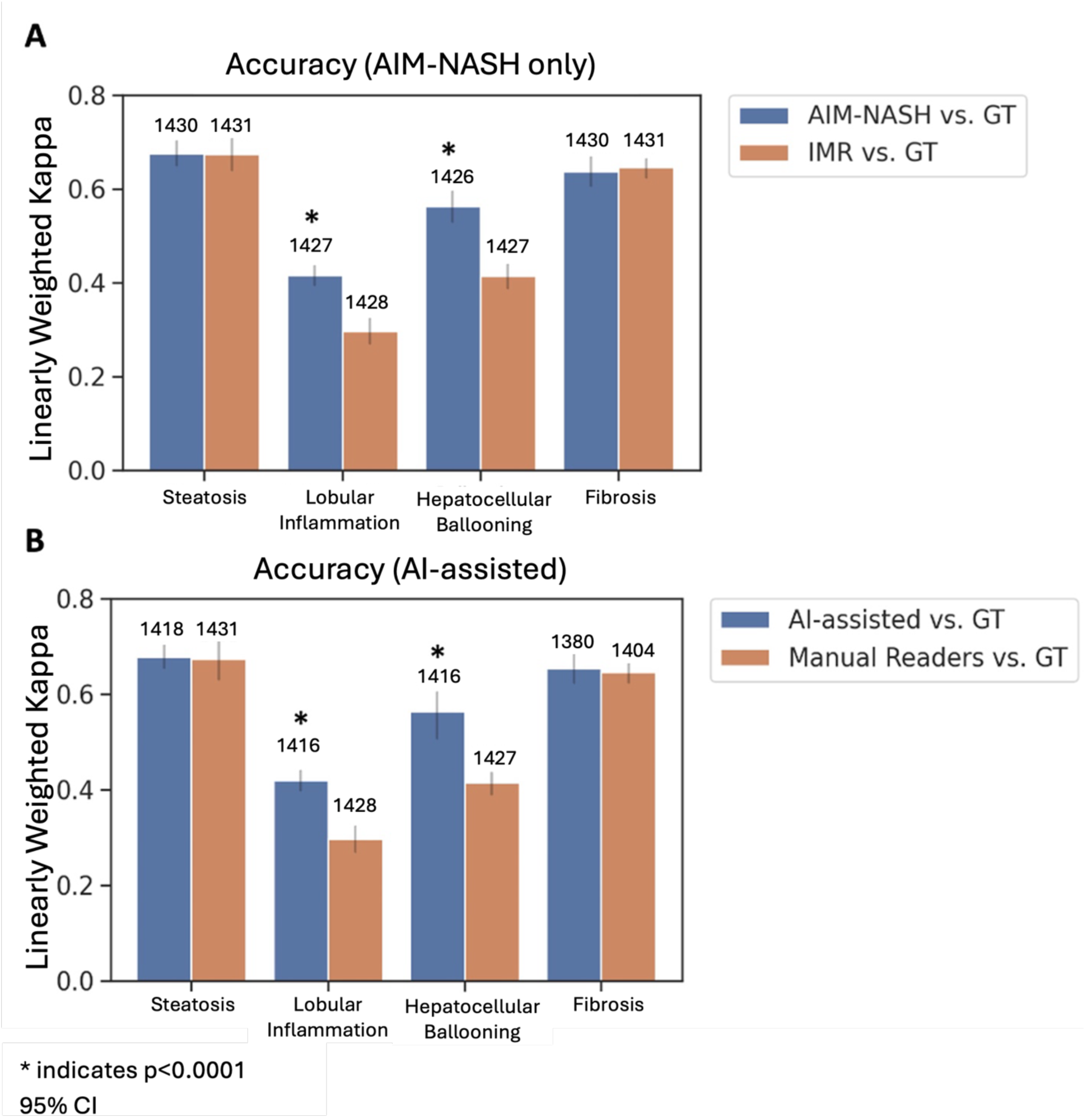
Accuracy concordance comparison of MASH histologic components. **(A)** AIM-NASH scores without pathologist review, **(B)** AI-assisted workflow. Dataset of 1,481 cases was used for clinical validation. **(A)** AIM-NASH scores without pathologists’ review workflow. **(B)** The AI-assisted workflow integrated pathologist review of the sample quality, staining, scanning adequacy, assessment of any additional findings and subsequent AIM-NASH scoring. Although pathologists could record minor disagreements with AIM-NASH scores, only major discrepancies (2 points or greater difference) permitted score alterations. * indicates statistical superiority. IMR – individual manual read; GT – ground truth

For AIM-NASH only **(Figure 3A)**, the difference in WK for AIM-NASH and GT compared to mean WK for IMR and GT for hepatocellular ballooning was 0.15 (95% CI of (0.11, 0.18); non-inferiority p<0.0001) and for lobular inflammation was 0.12 (95% CI of (0.08, 0.17); non-inferiority p<0.0001) with a p<0.0001 for superiority for both components. The difference in WK for AIM-NASH only and GT compared to WK of mean IMR and GT for steatosis was 0.01 (95% CI of (−0.02, 0.03); non-inferiority p<0.0001) and for fibrosis was −0.01 (95% CI of (−0.04, 0.02; non-inferiority p<0.0001). Steatosis and fibrosis met non-inferiority but did not achieve superiority.

For AI-assisted pathologist reading **(Figure 3B)**, the difference in WK for AI-assisted and GT compared to mean WK for IMR and GT for hepatocellular ballooning was 0.15 (95% CI of (0.11, 0.19); non-inferiority p<0.0001) and for lobular inflammation was 0.12 (95% CI of (0.08, 0.17); non-inferiority p<0.0001) with a p<0.0001 for superiority for both components. The difference in WK for AI-assisted and GT compared to mean WK for IMR and GT for steatosis was 0.01 (95% CI of (−0.02, 0.04); non-inferiority p<0.0001) and for fibrosis was 0.01 (95% CI of (−0.02, 0.03); non-inferiority p<0.0001). Steatosis and fibrosis met non-inferiority but did not achieve superiority. For all MASH score components WKs for AI-assisted and GT were in the ranges of published CRN pathologists WKs [8, 14].

For AI-assisted pathologist reading accuracy was higher for composite histologic scores as compared with IMRs **(Figure 4)**. The WKs for AI-assisted and GT and WKs for IMR and GT for fibrosis 2 and 3 (F2&F3) vs other were equivalent, with WK for AI-assisted and GT being slightly higher than WK for IMR and GT (0.57 vs 0.53, respectively; **Figure 4**). WKs for trial relevant enrollment criteria NAS ≥ 4 with ≥1 in each score category between AI-assisted and GT was significantly higher than the WK between IMR and GT ((0.63 vs 0.51, respectively, with a difference of 0.11 and 95% CI of (0.07 0.16)) and for MASH resolution (defined as hepatocellular ballooning score of 0, lobular inflammation score of 0 or 1 and any steatosis score) between AI-assisted and GT was also significantly higher than the WK between IMR and GT (0.54 vs 0.37, respectively, with a difference of 0.16 and 95% CI of (0.10, 0.22)) **(Figure 4**).

**Figure 4.**
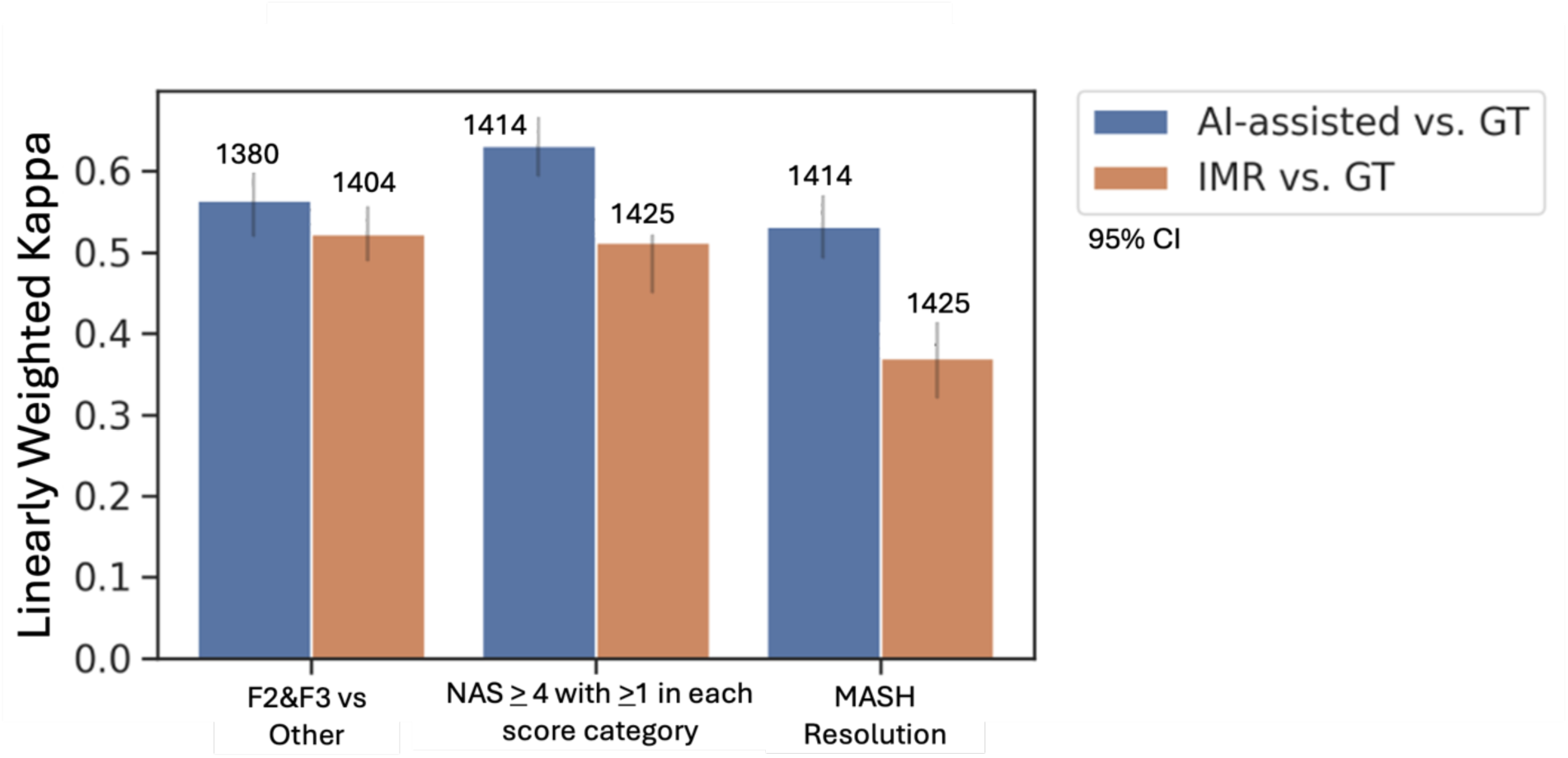
Comparisons for MASH aggregate component scores (F2&F3 vs other and NAS > 4 with >1 in each score category vs other) and MASH resolution. Aggregate components scores relevant to MASH clinical trial enrollment and endpoint assessment were calculated for AI-assisted vs ground truth (GT) and individual manual reader (IMR) vs GT. MASH resolution is defined as a hepatocellular ballooning score of 0, lobular inflammation 0 or 1, and any score for steatosis. AI-assisted reads for NAS≥4 with ≥1 in each component category and for MASH resolution were superior compared to independent manual reads.

For AI-assisted evaluation against a mode/median of a panel of pathologists, non-inferiority was met for all histologic components for agreement of AI-assisted reads with statistical ground truth reads, compared to the agreement between mode/median read scores derived from two different groups of pathologists (ground truth workflow in **Figure 1B and C**; results in **Figure 5**). For steatosis, the average WK for AI-assisted vs GT was 0.68 and for manual mode/median was 0.75, with a difference of –0.07; for lobular inflammation the WK for AI-assisted vs GT was 0.43 and for manual mode/median was 0.44, with a difference of –0.02; for hepatocellular ballooning the WK for AI-assisted vs GT was 0.56 and for manual mode/median was 0.53, with a difference of 0.04; and for fibrosis the WK for AI-assisted vs GT was 0.65 and for manual mode/median was 0.72, with a difference of –0.09.

**Figure 5.**
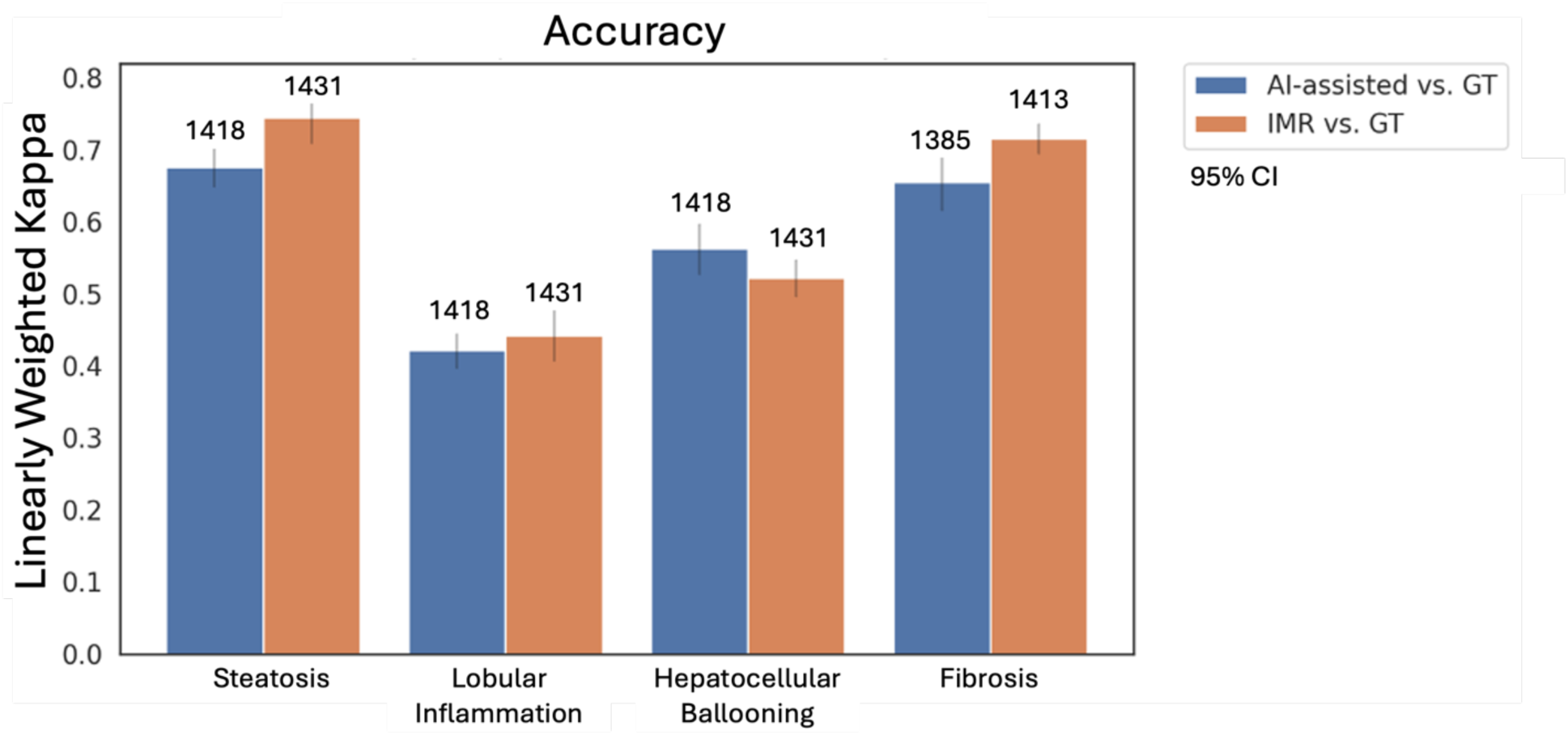
Weighted Kappa analysis for NASH components AI-assisted and mode/median panel comparisons. The same cohort of 1,481 cases utilized in analytical and clinical validation was utilized to determine accuracy of AI-assisted reads against 2 panels of readers. Statistical ground truth (GT) using mode/median scores instead of panel consensus and statistical mode/median individual manual read (IMR) panel derived from a minimum of 3 IMRs were determined.

## Discussion

AI-based tools have the potential to solve many of the issues around standardized, accurate and reproducible scoring, within and across trials. Multiple pathologists can assess biopsies on validated WSI viewers [24], both for sample adequacy, evaluability and for overall diagnosis and additional findings, while using AI tools to efficiently provide the accurate, standardized and consistent scores needed.

The AIM-NASH outputs have been validated according to their proposed use with representative trial datasets, both variable in disease activity, stain, scanning site, and drug candidate. Further, the overlays presented to the pathologist, identifying key areas that the model is predicting as artifact, steatosis, hepatocellular ballooning, lobular inflammation, and fibrosis, have been validated by multiple pathologist readers on a frames level, demonstrating they are highly sensitive and sufficiently specific in playing their role as a highlighter, to guide pathologist review, along with the associated model scores. These results demonstrate the precision of AIM-NASH in measuring each component of the CRN scoring system in liver biopsies from patients screened and/or enrolled in a MASH clinical trial.

Additionally, repeatability studies demonstrated superior performance of AIM-NASH when comparing to a performance goal of 85% as well as to relevant published manual intra-pathologist trial read agreements (steatosis 0.72, lobular inflammation 0.55, hepatocellular ballooning 0.70 and fibrosis 0.72) described in the literature [14]. AIM-NASH reproducibility across the three external laboratories, utilizing different operators and different Leica Aperio AT2 scanners, was higher for all MASH components than published inter-pathologist variability across expert MASH pathologists (0.63 for steatosis, 0.60 for lobular inflammation, 0.63 for hepatocellular ballooning and 0.51 for fibrosis) [14]. Further, the repeatability and reproducibility agreement achieved in this study with AIM-NASH was higher than the inter-pathologist agreement for independent manual reads.

Finally, the clinical validation study demonstrated that AIM-NASH consistently brought individual pathologists closer to ground truth reads for the histologic components historically most difficult to score (hepatocellular ballooning and lobular inflammation), while maintaining high levels of accuracy for steatosis and fibrosis. To evaluate AIM-NASH reads against a statistical consensus currently being used as a gold standard read during MASH trials, the agreement of AI-assisted reads with the median consensus of the ground truth reads, was compared to the agreement between two different median consensus groups (derived from IMR pathologist reads and GT pathologist reads) in the same non-inferiority analysis used in the primary endpoint for accuracy. AI-assisted reads achieved non-inferiority for every histologic component score in this analysis, and AI-assisted read agreement with median ground truth for hepatocellular ballooning was higher than that for median IMR agreement with median ground truth. For steatosis, although the two manual median groups mean agreement with each other was higher than that for AIM-NASH vs. median ground truth, AI-assisted reads were still within the non-inferiority margin and should be interpreted as such. Additionally, accuracy and reproducibility are inter-connected in the MASH trial context of use for assessment of primary endpoints, and AIM-NASH provides for a more reliable, reproducible read across all components. Furthermore, the gold standard is still subject to enrollment bias and lack of standardization, as demonstrated by the Kappas achieved by the median IMR vs. median ground truth in this study (**Figure 5**) and supported by the findings from Sanyal et al., [12] which evaluated agreement between two gold standard panel reads. Finally, the achievement of non-inferiority by AIM-NASH for accuracy compared to a gold standard read across a robust clinical validation dataset, provides strong evidence that AIM-NASH agrees with two consensus groups as well as they agree with each other, and, therefore, could replace the current gold standard consensus read approach for trials while enabling a more standardized, less biased approach to accurately enrolling and determining change in score over time for primary histologic endpoints.

The sum of the ordinal scores for steatosis, lobular inflammation, and hepatocellular ballooning (NAS) being greater than or equal to 4 (NAS≥4) is one of the main indicators for a probable MASH diagnosis, as well as commonly being a requirement for trial inclusion. Additionally, one component of the composite endpoint is MASH resolution, defined as a hepatocellular ballooning score of 0, lobular inflammation 0 or 1, and any score for steatosis. AI-assisted reads for NAS≥4 with ≥1 in each component category and for MASH resolution were superior compared to independent manual reads. This is an important indicator that AIM-NASH can be a powerful tool in increasing and standardizing key aspects of trial scoring for enrollment and for FDA and EMA recommended endpoints.

In addition, the AIM-NASH algorithm alone has also demonstrated to either recapitulate or demonstrate that primary efficacy results were met across several trials and drug candidates (semaglutide, pegbelfermin, resmetirom [25–28]). In a phase 2b study for pegbelfermin AIM-NASH revealed a statistically significant difference in the proportion of primary endpoint responders in treatment vs. placebo groups, whereas the central pathologist scoring did not reveal a statistically significant difference [26]. In a phase 2b study for resmetirom, all endpoints met via both individual manual readers were also met by AIM-NASH [27]. In the phase 3 study for resmetirom, for both MASH resolution and fibrosis improvement endpoint, the percentage of patients that responded were comparable when assessed by AIM-NASH or manual pathology assessment [28]. Lastly, in a cirrhotic patient population from another phase 2 study for semaglutide, a numerically higher proportion of patients was seen across both assessment methods (AIM-NASH and manual reads) for semaglutide vs placebo for inflammation, steatosis and ballooning from baseline to week 48. Additionally, a lower placebo effect response was observed with AIM-NASH compared to manual reads [25]. This supportive evidence, along with the accuracy of AIM-NASH alone and AI-assisted evidence demonstrates the robust nature of the AIM-NASH across a wide range of disease activity and in the phase of drug treatments.

As the samples for this study were sourced from completed clinical trials with a wide range of sample quality and the reads were performed retrospectively, the limitations of the study include the inability of the pathologists to request a re-stain or a rescan of samples where they thought the sample was not of sufficient quality. This could have led to higher rates of samples being deemed inadequate or non-evaluable for scoring, as in a clinical trial setting these samples could be re-stained or rescanned. However, these cases represented less than 4% of all clinical validation cases. Additionally, although the dataset was large and robust, new trial populations and/or drug candidates with novel mechanisms of actions not encountered here could potentially present a challenge to the algorithm in its current state.This highlights the importance of the pathologist evaluation and quality control of the algorithm results, and performance monitoring will be utilized to indicate where there may be room for future improvement through additional training.

Together, the above data supports the use of AIM-NASH by pathologists in trials and can play a significant role in resolving the accuracy and precision gaps in MASH assessment, while guiding pathologists in an efficient evaluation to result in a standardized and reproducible score within and across trials. This in turn could significantly benefit MASH patients in helping to bring truly effective therapies to market.

## Online Methods

### Datasets and Study Oversight

The analysis utilized existing de-identified glass slides and WSIs derived from liver biopsies procured during four MASH clinical trials, including three phase 2 trials and one phase 3 trial (screen failures and enrolled population from Intercept Pharmaceuticals REGENERATE trial NCT02548351, enrolled population from Bristol Myers Squibb FALCON 1 trial NCT03486899 and FALCON 2 trial NCT03486912 and enrolled population from Novo Nordisk Semaglutide trial NCT02970942). These data encompassed a broad spectrum of disease manifestations, captured both screened and enrolled participants, and mirrored the variances observed in the MASH clinical trial population. Sample collection varied, encompassing historical and study biopsies, with staining procedures executed across multiple sites. The research was granted expedited approval by the WCG Institutional Review Board (IRB00000533).

### AIM-NASH Development

AIM-NASH was trained using 103,579 pathologist-provided annotations of 6235 hematoxylin and eosin (H&E) and 6223 Masson’s trichrome WSIs from 6 completed phase 2b and phase 3 MASH clinical trials. For every WSI, AIM-NASH employs a sequential approach where convolutional neural networks produce tissue overlays containing colorized predictions of segmentation, signifying various histologic features.

Additionally, slide-level quantifications of the proportionate area of each feature are generated. Simultaneously, graph neural networks predict an ordinal MASH CRN grade or stage for each histologic feature. The development of AIM-NASH is further described in Iyer et al 2023 [22].

### Overlay Validation Analyses

In order to assess the accuracy of the heatmap overlays generated by the AIM-NASH model to enable efficient review of key histologic features considered by the algorithm, up to 160 500 x 500-micron sized frames for each feature (steatosis, lobular inflammation, hepatocellular ballooning, fibrosis, H&E artifact and trichrome artifact) were selected to represent a wide range of each histology and commonly encountered artifacts (e.g., tissue folds, stain pooling, scanning blur). Only usable tissue is considered in predicting scores. These overlays are intended to facilitate the pathologist’s review in the AIM-NASH scoring workflow and therefore, were designed with preference for sensitivity. The enrolling pathologist estimated the amount of each feature in each frame on images with no overlays. Three board-certified expert hepatopathologists were provided with the enrolled frames from both H&E and trichrome slides. The pathologists were asked specific questions for each frame to determine to what extent the overlay may or may not be under- or overestimating a given feature, defined as true positive (TP) and false positive (FP) success rates. Overlay performance was considered acceptable if TP success rate and FP success rate were greater than or equal to 85%.

Frames from 222 WSIs were enrolled. Overall, 312 unique H&E frames and 249 trichrome frames were enrolled from 3 clinical trials (both baseline and follow-up time points from placebo and treatment groups).

### Repeatability and Reproducibility Analyses

For the assessment of AIM-NASH’s reproducibility, we incorporated glass slides from two completed phase 2 trials (one non-cirrhotic and one cirrhotic) and a phase 3 MASH trial. To gauge inter-day repeatability, 150 cases, each comprising a H&E and a trichrome slide, were repeatedly scanned using the same Leica Aperio AT2 scanner at 40x magnification across three non-sequential days. For inter-site reproducibility assessment, identical cases were singularly scanned at 3 distinct labs by different operators using separate AT2 scanners. Reproducibility and repeatability were deemed acceptable when mean pairwise agreement rates consistently matched or surpassed 85%. No pathologist review of AIM-NASH scores was incorporated in repeatability and reproducibility studies.

### Establishment of Ground Truth

Ground truth (GT), defined as the presumed accurate diagnosis, was ascertained by dual panels of hepatic pathologists. Each panel consisted of two main reader pathologists and an auxiliary tie-breaker pathologist. Discrepancies in scoring among the primary readers prompted the intervention of the tiebreaker pathologist, who was blinded to initial assessments. When the tiebreaker’s scoring diverged from both primary readers, a panel discussion was convened for consensus, with the tiebreaker’s score being decisive in rare cases of continued disagreement. Overall, five distinct pathologists contributed to establishing the ground truth (**Figure 1B**).

### Analytical Validation Protocol

For analytical validation, 1,481 cases extracted from two finalized phase 2 trials and select cases from a phase 3 trial, representing 3 different drug candidates with unique mechanisms of action (semaglutide, pegbelfermin, resmetirom), were evaluated in comparison to GT and individual manual reader (IMR). Cases from the phase 3 trial were selected to match the original trial enrolled population (baseline and follow-up timepoints) and included screen failures. Each case underwent scanning via a Leica Aperio AT2 scanner at 40x magnification. Notably, this phase excluded pathologist review of resultant scores.

### Clinical Validation Protocol

The same cohort of 1,481 cases incorporated in the analytical validation phase was used for clinical validation. This phase aimed to ascertain AIM-NASH’s capability to bolster pathologists’ accuracy in MASH diagnosis in a therapeutic trial context. The AI-assisted workflow integrated pathologist review of the sample quality, staining, scanning adequacy, assessment of any additional findings and subsequent AIM-NASH scoring. Although pathologists could record minor disagreements with AIM-NASH scores, only major discrepancies (2 points or greater difference) permitted score alterations **(Figure 1A**).

### Panel comparison

The same cohort of 1,481 cases utilized in analytical and clinical validation was utilized to determine accuracy of AI-assisted reads against 2 panels of readers. Statistical ground truth using mode/median scores instead of panel consensus (**Figure 1C**) and statistical median derived from a minimum of 3 IMRs were determined.

### Statistical Analysis

Both analytical and clinical validation phases were designed to initially assess AIM-NASH’s non-inferiority to manual scoring. Upon confirmation of non-inferiority, its accuracy was further assessed for superiority. Non-inferiority was established when the difference between AIM-NASH Cicchetti-Allison kappa with the GT exceeded a non-inferiority margin of −0.1 compared to the independent manual read (IMR) weighted Kappa (WK) with the GT for each MASH component (Bootstrap percentile p < 0.025). Linearly WK was utilized, as pairwise comparisons are used to determine the level of agreement and using this metric, agreement between raters adjusting for the agreement that might occur by chance could be computed. The linear weights, in this case, penalize disagreement due to distant scores (e.g. 3 vs. 1) more compared to that between closer ordinal scores (e.g. 2 vs. 1).

## Data Availability

The glass slides and WSIs used in these validation studies are from existing clinical trials and the authors had access to these during the study in accordance with the relevant license agreements. Due to the nature of the source data, it is not currently publicly available.

## Code Availability

Not all original code can be made publicly available. The code for cell- and tissue-type model training, inference, and feature extractions are not disclosed. To safeguard PathAI’s intellectual property, access requests for such code will not be considered. The source code for all downstream data analyses and figure generation in this work are publicly available and can be downloaded from GitHub: https://github.com/Path-AI/NASH_DDT_Manuscript

## Acknowledgements

We wish to thank all the patients and family members that participated in the clinical trials. We would also like to thank all the pathologists who participated in the study. This study was funded by PathAI, Inc.

## Supplementary Information

**Table S1.** Frame Distribution based on Slide Level Score for Overlay Validation.

| Feature | Score | (n/N) | % |
| --- | --- | --- | --- |
| Hepatocellular ballooning | 0 | (13/86) | 15.1 |
|  | 1 | (36/86) | 41.9 |
|  | 2 | (37/86) | 43.0 |
| Lobular inflammation | 0 | (2/87) | 2.3 |
|  | 1 | (46/87) | 52.9 |
|  | 2 | (31/87) | 35.6 |
|  | 3 | (8/87) | 9.2 |
| Steatosis | 0 | (5/87) | 5.8 |
|  | 1 | (31/87) | 35.6 |
|  | 2 | (31/87) | 35.6 |
|  | 3 | (20/87) | 23.0 |
| Fibrosis | 0 | (1/79) | 1.27 |
|  | 1 | (16/79) | 20.3 |
|  | 2 | (21/79) | 26.6 |
|  | 3 | (30/79) | 38.0 |
|  | 4 | (11/79) | 13.9 |

Note: N<160 since multiple frames can be selected from a slide

**Table S2:** Frame Distribution based on Frames Level Score for Overlay Validation.

| Feature | Score Category | (n/N) | % |
| --- | --- | --- | --- |
| H&E Artifact | None | (20/160) | 12.5 |
|  | Present | (140/160) | 87.5 |
| Hepatocellular ballooning | None | (16/160) | 10.0 |
|  | 1-Few | (72/160) | 45.0 |
|  | Frequent | (72/160) | 45.0 |
| Lobular inflammation | None | (11/160) | 6.88 |
|  | 1 | (50/160) | 31.3 |
|  | 2-4 | (50/160) | 31.3 |
|  | >4 | (49/160) | 30.6 |
| Steatosis | None | (10/160) | 6.25 |
|  | Low | (50/160) | 31.3 |
|  | Medium | (50/160) | 31.3 |
|  | High | (50/160) | 31.3 |
| Trichrome Artifact | None | (22/160) | 13.8 |
|  | Present | (138/160) | 86.3 |
| Fibrosis | None | (10/160) | 6.25 |
|  | Low | (53/160) | 33.1 |
|  | Medium | (50/160) | 31.3 |
|  | High | (47/160) | 29.4 |

**Table S3:** Presence of Feature per Pathologist for Overlay Validation.

| Feature | Pathologist | Presence | (n/N) | % |
| --- | --- | --- | --- | --- |
| H&E Artifact | A | Yes | (141/160) | 88.1 |
|  | B | Yes | (135/160) | 84.4 |
|  | C | Yes | (143/160) | 89.4 |
| Hepatocellular ballooning | A | Yes | (92/160) | 57.5 |
|  | B | Yes | (71/160) | 44.4 |
|  | C | Yes | (111/160) | 69.4 |
| Lobular inflammation | A | Yes | (132/160) | 82.5 |
|  | B | Yes | (132/160) | 82.5 |
|  | C | Yes | (155/160) | 96.9 |
| Steatosis | A | Yes | (159/160) | 99.4 |
|  | B | Yes | (158/160) | 98.8 |
|  | C | Yes | (159/160) | 99.4 |
| Trichrome Artifact | A | Yes | (114/160) | 71.3 |
|  | B | Yes | (124/160) | 77.5 |
|  | C | Yes | (149/160) | 93.1 |
| Fibrosis | A | Yes | (151/160) | 94.4 |
|  | B | Yes | (150/160) | 93.8 |
|  | C | Yes | (153/160) | 95.6 |

**Table S4.**
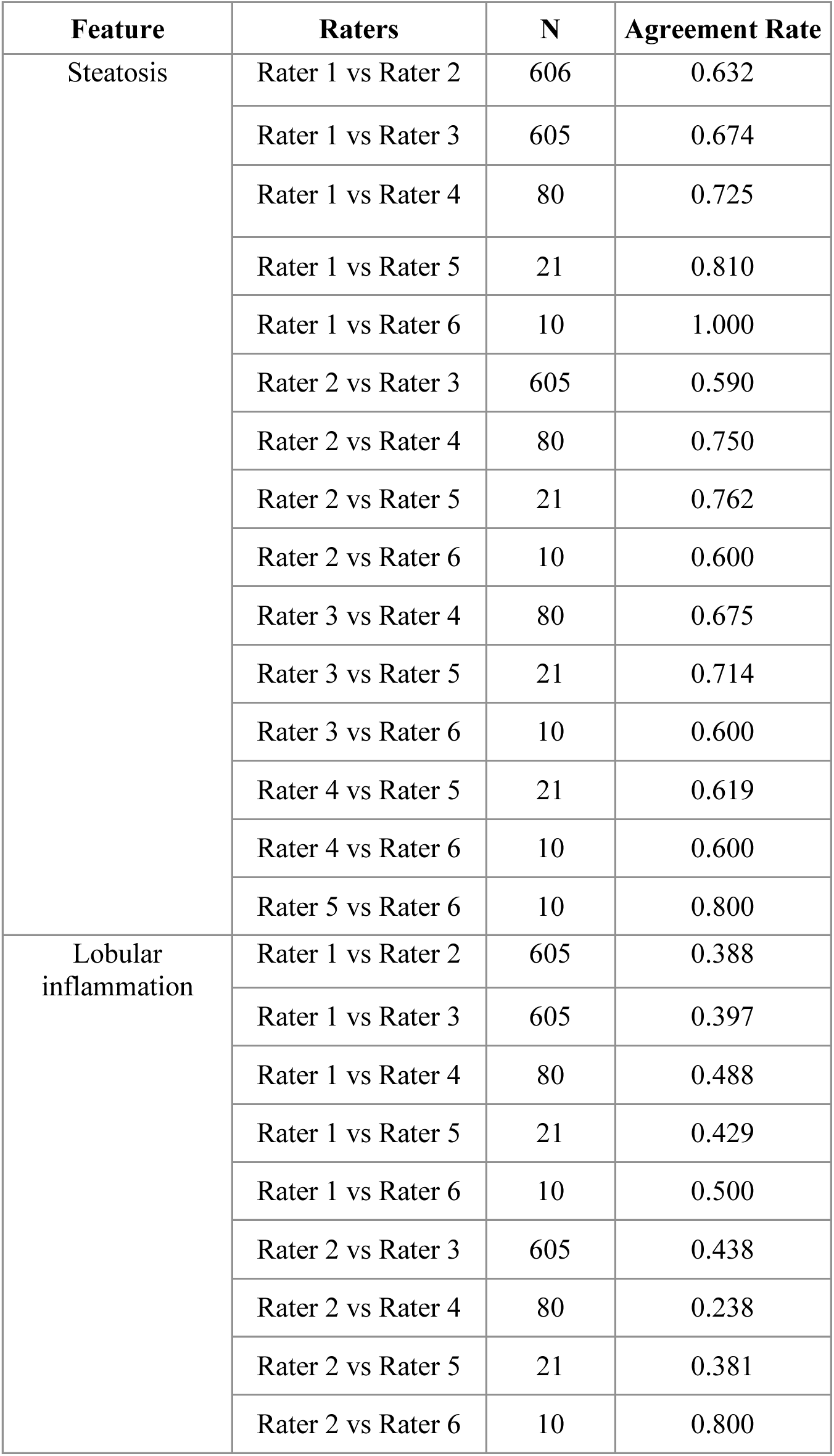
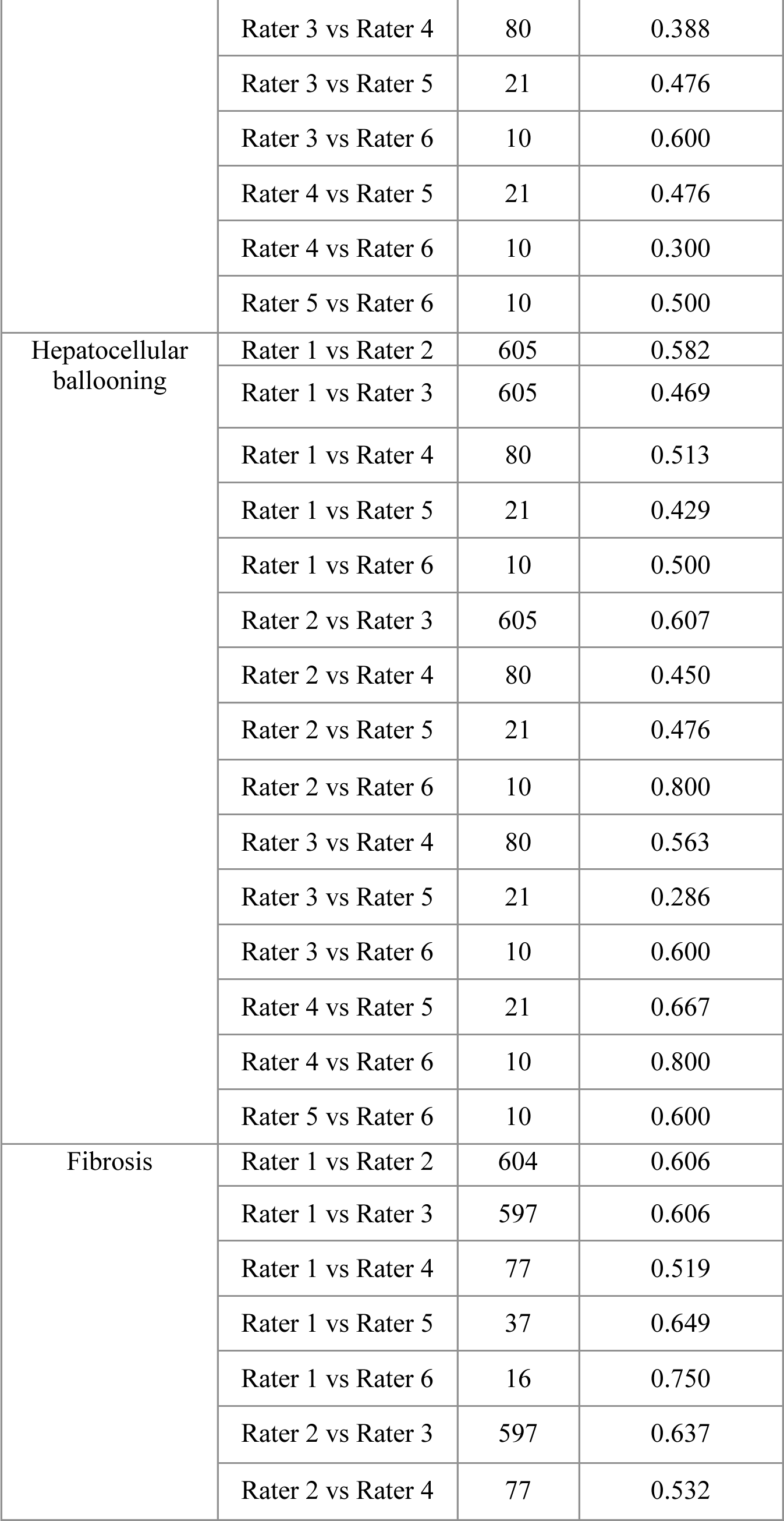
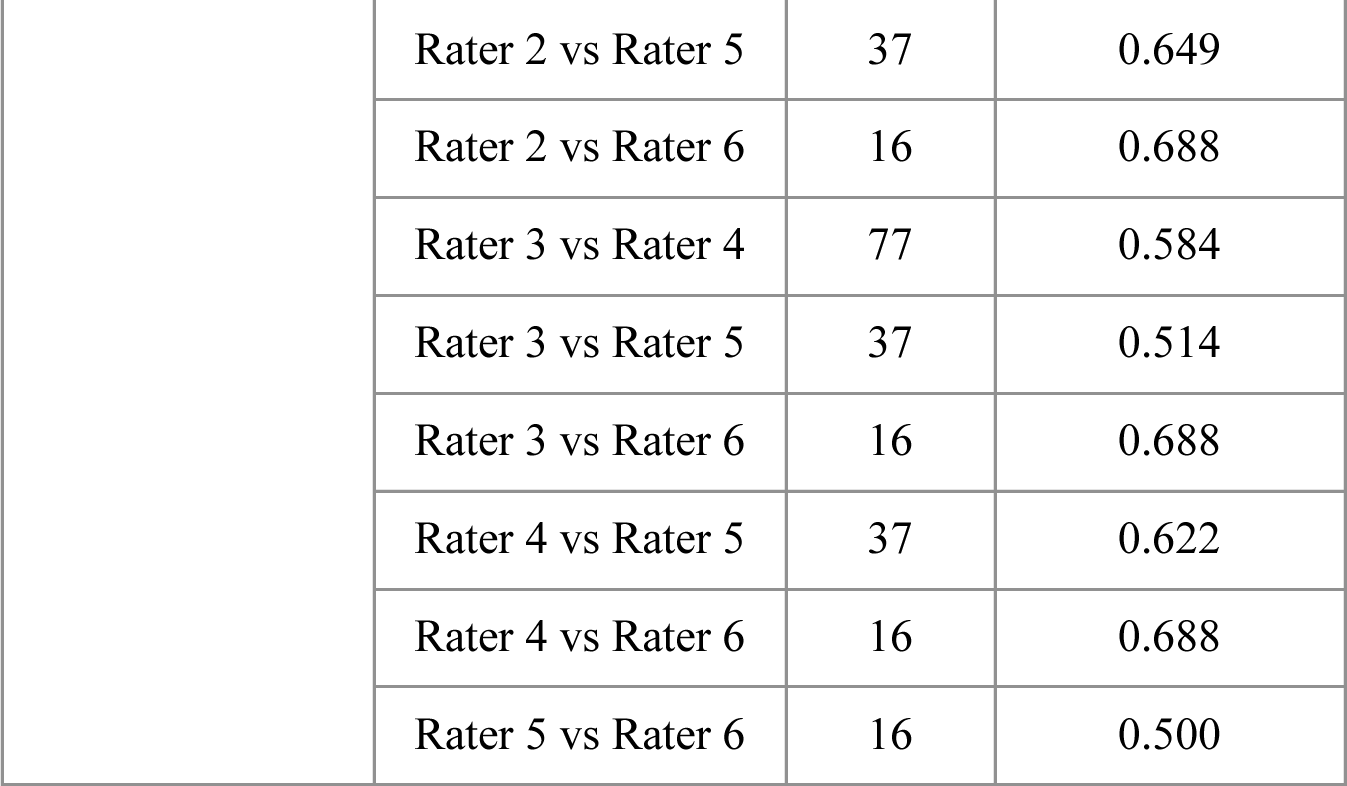
Pairwise inter-reader agreement rates.

## References

[1] Rinella ME, Lazarus J V., Ratziu V, et al. A multisociety Delphi consensus statement on new fatty liver disease nomenclature. Journal of Hepatology 2023; 79: 1542–1556.

[2] Younossi ZM, Koenig AB, Abdelatif D, et al. Global epidemiology of nonalcoholic fatty liver disease—Meta-analytic assessment of prevalence, incidence, and outcomes. Hepatology 2016; 64: 73–84.

[3] Noureddin M, Vipani A, Bresee C, et al. NASH Leading Cause of Liver Transplant in Women: Updated Analysis of Indications For Liver Transplant and Ethnic and Gender Variances. American Journal of Gastroenterology 2018; 113: 1649–1659.

[4] Friedman SL, Neuschwander-Tetri BA, Rinella M, et al. Mechanisms of NAFLD development and therapeutic strategies. Nature Medicine 2018; 24: 908–922.

[5] FDA-NIH Biomarker Working Group. BEST (Biomarkers, EndpointS, and other Tools) Resource [Internet]. 2016.

[6] FDA. Nonalcoholic Steatohepatitis with Compensated Cirrhosis: Developing Drugs for Treatment Guidance for Industry - Draft.

[7] FDA. Noncirrhotic Nonalcoholic Steatohepatitis With Liver Fibrosis: Developing Drugs for Treatment Guidance for Industry.

[8] Brunt EM, Kleiner DE, Wilson LA, et al. Improvements in Histologic Features and Diagnosis Associated With Improvement in Fibrosis in Nonalcoholic Steatohepatitis: Results From the Nonalcoholic Steatohepatitis Clinical Research Network Treatment Trials. Hepatology 2019; 70: 522–531.

[9] Tong X fei, Wang Q yi, Zhao X yan, et al. Histological assessment based on liver biopsy: the value and challenges in NASH drug development. Acta Pharmacologica Sinica 2022; 43: 1200–1209.

[10] EMA. Reflection paper on regulatory requirements for the development of medicinal products for chronic non-infectious liver diseases (PBC, PSC, NASH) (EMA/CHMP/299976/2018). European Medicines Agency; 44.

[11] Harrison SA, Bedossa P, Guy CD, et al. A Phase 3, Randomized, Controlled Trial of Resmetirom in NASH with Liver Fibrosis. New England Journal of Medicine; 390. Epub ahead of print 2024. DOI: 10.1056/nejmoa2309000.

[12] Sanyal AJ, Loomba R, Anstee QM, et al. Utility of pathologist panels for achieving consensus in NASH histologic scoring in clinical trials: Data from a phase 3 study. Hepatol Commun; 8. Epub ahead of print January 2024. DOI: 10.1097/hc9.0000000000000325.

[13] Kleiner DE, Brunt EM, Van Natta M, et al. Design and validation of a histological scoring system for nonalcoholic fatty liver disease. Hepatology 2005; 41: 1313–1325.

[14] Davison BA, Harrison SA, Cotter G, et al. Suboptimal reliability of liver biopsy evaluation has implications for randomized clinical trials. J Hepatol 2020; 73: 1322–1332.

[15] Merriman RB, Ferrell LD, Patti MG, et al. Correlation of paired liver biopsies in morbidly obese patients with suspected nonalcoholic fatty liver disease. Hepatology 2006; 44: 874–880.

[16] Juluri R, Vuppalanchi R, Olson J, et al. Generalizability of the nonalcoholic steatohepatitis clinical research network histologic scoring system for nonalcoholic fatty liver disease. J Clin Gastroenterol 2011; 45: 55–58.

[17] Pavlides M, Birks J, Fryer E, et al. Interobserver variability in histologic evaluation of liver fibrosis using categorical and quantitative scores. Am J Clin Pathol 2017; 147: 364–369.

[18] Sanyal A, Loomba R, Anstee Q, et al. Minimizing Variability and Increasing Concordance for NASH Histological Scoring in NASH Clinical Trials. In: American Association for the Study of Liver Diseases. 2021.

[19] Harrison SA, Alkhouri N, Davison BA, et al. Insulin sensitizer MSDC-0602K in non-alcoholic steatohepatitis: A randomized, double-blind, placebo-controlled phase IIb study. J Hepatol 2020; 72: 613–626.

[20] Muehlematter UJ, Daniore P, Vokinger KN. Approval of artificial intelligence and machine learning-based medical devices in the USA and Europe (2015–20): a comparative analysis. The Lancet Digital Health 2021; 3: e195–e203.

[21] Perincheri S, Levi AW, Celli R, et al. An independent assessment of an artificial intelligence system for prostate cancer detection shows strong diagnostic accuracy. Modern Pathology 2021; 34: 1588–1595.

[22] Iyer JS, Pokkalla H, Biddle-Snead C, et al. AI-based histologic scoring enables automated and reproducible assessment of enrollment criteria and endpoints in NASH clinical trials. medRxiv 2023; 2023.04.20.23288534.

[23] Brunt EM, Clouston AD, Goodman Z, et al. Complexity of ballooned hepatocyte feature recognition: Defining a training atlas for artificial intelligence-based imaging in NAFLD. J Hepatol 2022; 76: 1030–1041.

[24] Pulaski H, Mehta SS, Manigat LC, et al. Validation of Digital Pathology Platform for Metabolic-Associated Steatohepatitis for Clinical Trials. medRxiv 2023; 2023.09.01.23294940.

[25] Loomba R, Cejvanovic V, Iyer J, et al. Comparison of the effects of semaglutide on liver histology in patients with non-alcoholic steatohepatitis cirrhosis between machine learning model assessment and pathologist evaluation. In: American Association for the Study of Liver Diseases. 2022.

[26] Shevell D, Brown E, Minnich A, et al. Comparison of manual vs machine learning approaches to liver biopsy scoring for NASH and fibrosis: a post hoc analysis of the FALCON 1 study. In: American Association for the Study of Liver Diseases. 2021.

[27] Harrison S, Iyer J, Biddle-Snead C, et al. Retrospective AI-based measurement of NASH histology (AIMNASH) analysis of biopsies from Phase 2 study of Resmetirom confirms significant treatment-induced changes in histologic features of non-alcoholic steatohepatitis. In: European Association for the Study of the Liver. London, UK, p. S711.

[28] Iyer J, Bedossa P, Guy C, et al. Artificial Intelligence-based Measurement of NASH Histology (AIM-NASH) recapitulates primary results from Phase 3 study of resmetirom for treatment of NASH/MASH with liver fibrosis. In: American Association for the Study of Liver Diseases. Boston, MA, 2023, p. 24.

